# A Tempo-geographic Analysis of Global COVID-19 Epidemic Outside of China

**DOI:** 10.1101/2020.03.20.20039602

**Authors:** Huipeng Liao, Gifty Marley, Yafei Si, Zaisheng Wang, Yewei Xie, Cheng Wang, Weiming Tang

## Abstract

**Background:** Understanding the global epidemic trends, geographic distribution, and transmission patterns of COVID-19 contribute to providing timely information for the global response of the epidemic. This study aims to understand the global pandemic geospatial patterns and trends and identify new epicenters requiring urgent attention.

**Methods:** Data on COVID-19 between 31^st^ Dec. 2019 and 14^th^ Mar. 2020 was included. The epidemic trend was analyzed using joinpoint regressions; the growth of affected countries was by descriptive analysis; and the global distribution and transmission trend by spatial analysis. **Findings:** The number of new cases in the regions outside of China slowly increased before 24^th^ Feb. and rapidly accelerated after 24^th^ Feb. Compared to China, other affected countries experienced a longer duration of a slow increase at the early stage and rapid growth at the latter stages. The first apparent increase in the number of affected countries occurred from 23^rd^ Jan to 1^st^ Feb, and the second apparent increase started from 25^th^ Feb. The fist COVID-19 cases reported by countries from 28^th^ Feb. were mainly imported from Europe. The geographic distribution changed from single-center (13^th^ Jan. - 20^th^ Feb.) to multi-centers pattern (20^th^ Feb. – 14^th^ Mar.). More countries were affected with COVID-19 and developed local transmission.

**Interpretation:** The joinpoint regression and geospatial analysis indicated a multi-center pandemic of COVID-19. Strategies to prevent the new multiple centers as well as prevent ongoing transmission are needed.

**Funding:** NIH.

## Introduction

After the outbreak of COVID-19 in China, and after the epidemic turning better in China, the world outside of China started to experience an ongoing and worsening epidemic. According to the report of the World Health Organization (WHO), by 14^th^ Mar., 2020 a total of 142,539 cases have been reported in 136 countries and territories, and 9,769 new cases were reported on the single day of 14^th^ Mar.[1] Italy, Iran, and South Korea aside China have the highest number of confirmed cases (17,660; 11,364; 8086) as of 14^th^ Mar., 2020.[1] In response, highly affected countries have adopted innovative curbing interventions such as whole city lockdowns, the establishment of drive-through screening centers and banning large social gatherings amongst others to locally contain the virus and prevent further new cases.[2,3] Affected and unaffected countries following WHO guidelines are developing or have commenced implementation of country-specific prevention, early case detection as well as quarantine and treatment policies.[4] Some countries have closed borders and/or suspended flight commutation with major affected countries including South Korea and Iran regardless of the WHO cautions.[5,6] Such drastic measures are very essential in the pandemic control, but may also negatively affect the global economy if maintained over a longer duration as countries like China and South Korea contribute largely to global industrial production and provision of goods and services.[7,8]

Although some low-middle income countries (LMICs) in Africa remained unaffected for a while, 15 African countries and territories have reported a total of 73 confirmed cases as of 14^th^ Mar., 2020.[1] This furthers concerns of a pandemic as under-resourced LMICs in the region lack adequate infrastructures to successfully contain a breakout and are dependent on external sources for medical goods procurement.[4] Therefore, further spread of the virus threatens shortage to medical supplies and further outbreaks in LMICs could lead to high COVID-19 related mortality.[9] As such prompt responses and global strategic alliances are required immediately to prevent further outbreaks and avert the threatening pandemic.

As more countries get affected and the reported number of cases outside China keeps growing with each passing day, it is important to study the global disease outbreak, with especial focus on disease transmission, development trend, and new epicenters. Obtaining and analyzing this data will not only expand current knowledge on the disease global trends but also inform strategic planning and alliance for prompt global containment of the disease and prevention of further outbreaks.

This study aims to understand the global disease pandemic geo-spatial pattern, epidemic trends and identify new and emerging epicenters requiring urgent attention for decision making.

## Methods

The research studied the COVID-19 pandemic outbreak at the aspects of the global epidemic trend, geographic transmission patterns, and the changes in disease distribution from 13^th^ Jan. to 14^th^ Mar. 2020.

### Data collection and processing

The disease data of China were collected from China’s national health commission. The disease data outside of China were mainly collected from WHO daily COVID-19 situation reports,[1] except for the disease data before 19^th^ Jan. and the data about the origins of the first cases reported by each country and territory, which were collected from the news in terms of limited open-source data of COVID-19. The spatial data were derived from GADM version 3.6.[10] All the data were publicly available.

### Data Analysis

Statistical analyses were done using Joinpoint Trend Analysis version 4□7□0□0 Software (National Cancer Institute, USA) and R Studio software (R Core Team, 2016). The geographic outputs were generated by ArcGIS 10□2 software (Esri Inc, Redlands, California).

The pandemic disease outbreak was studied and analyzed through the epidemic trend over time by joinpoint regressions, the growth of infected countries and the number of infections over time, the global transmission routes of first reported cases in countries, and the geographic distribution over time by dynamic maps.

## Results

As of 14^th^ Mar., SRAS-CoV-2 has affected 134 countries and territories outside of China with 61,518 cases reported.

### The epidemic trend of COVID-19 outside of China

Globally, the joinpoint regression models showed that both the number of new cases and the number of cumulative cases were in rapid growth (Figure 1A and 1B). The trend of new cases was on the first stage before 24^th^ Feb. (slope = 4□4), where the number increased slightly and was smaller than 300. The number of new cases was then rapidly accelerated at the second to fourth stage from 24^th^ Feb. – 4^th^ Mar. (slope = 207□6), 4^th^ – 11^th^ Mar. (slope = 384□9), and 11^th^ – 14^th^ Mar. (slope = 1622□5). Compared to China (Figure 1C), the world outside of China stayed at the first stage of slow growth for a long period (43 days from 13^th^ Jan. – 24^th^ Feb. 2020; China: 24 days from 31^st^ Dec. 2019 to 23^rd^ Jan. 2020). The number of new cases accelerated for 15 days in China and gradually decreased after 6^th^ Feb., except for 12^th^ Feb(the diagnosis criteria changed by including clinically diagnosed cases, to facilitate treatment). The acceleration of new cases in the world outside of China started from 24^th^ Feb. and lasted for 20 days, which was already longer than that in China. However, a decreasing trend in the number of new cases outside of China was not yet observed as at 14^th^ Mar., and thus this stage might last longer, while China already achieved this on 12^th^ Feb.

**Figure 1.**
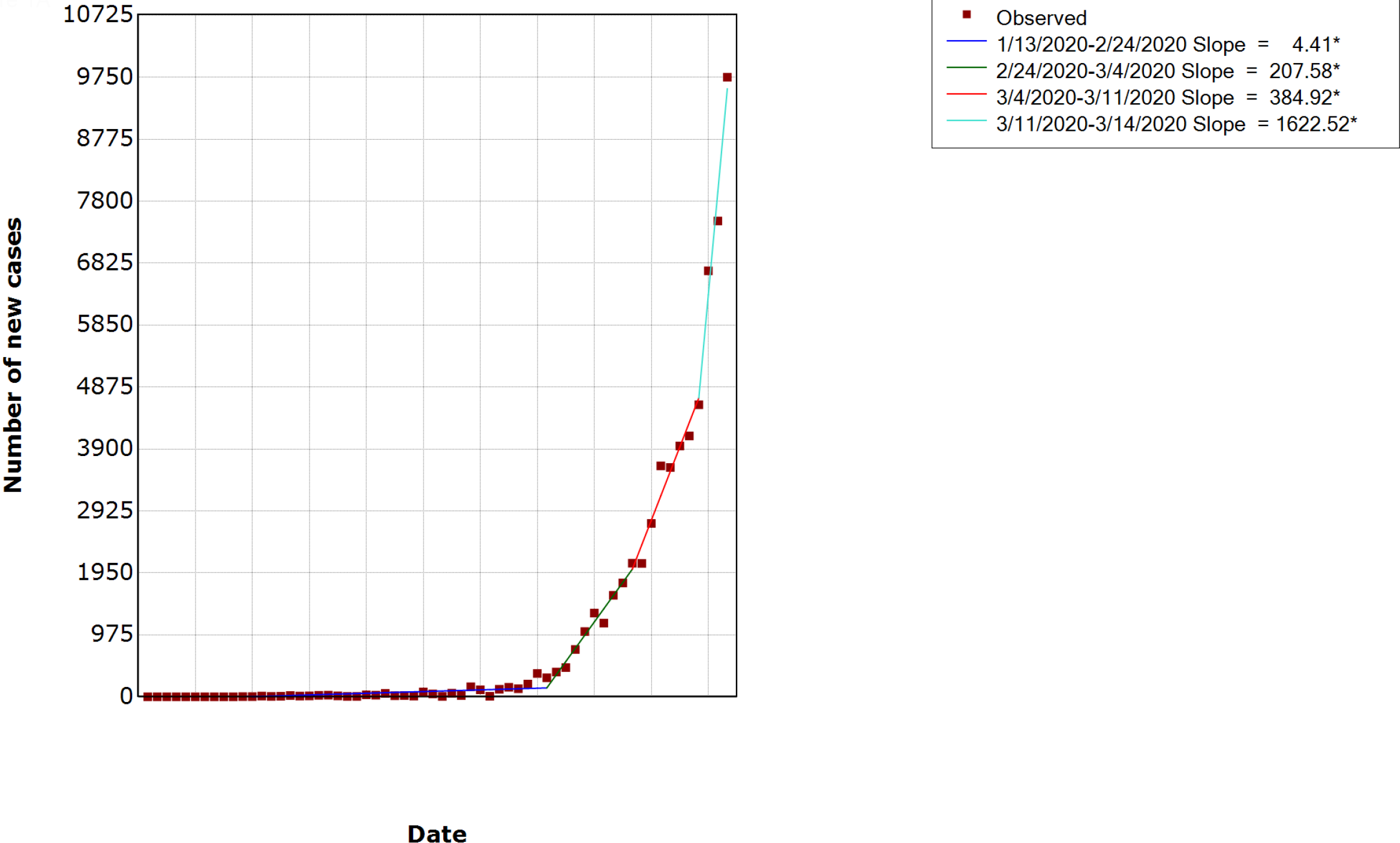
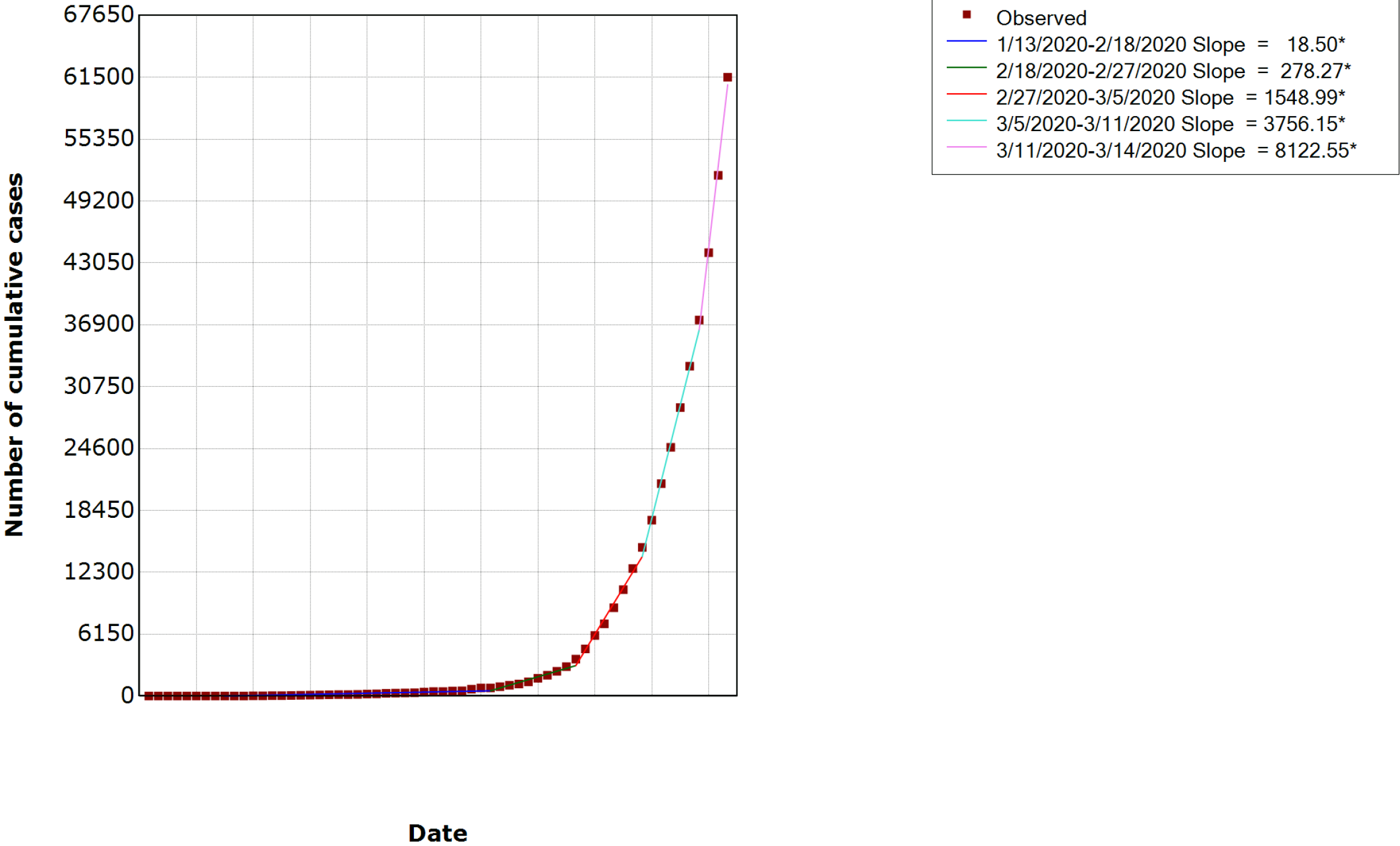
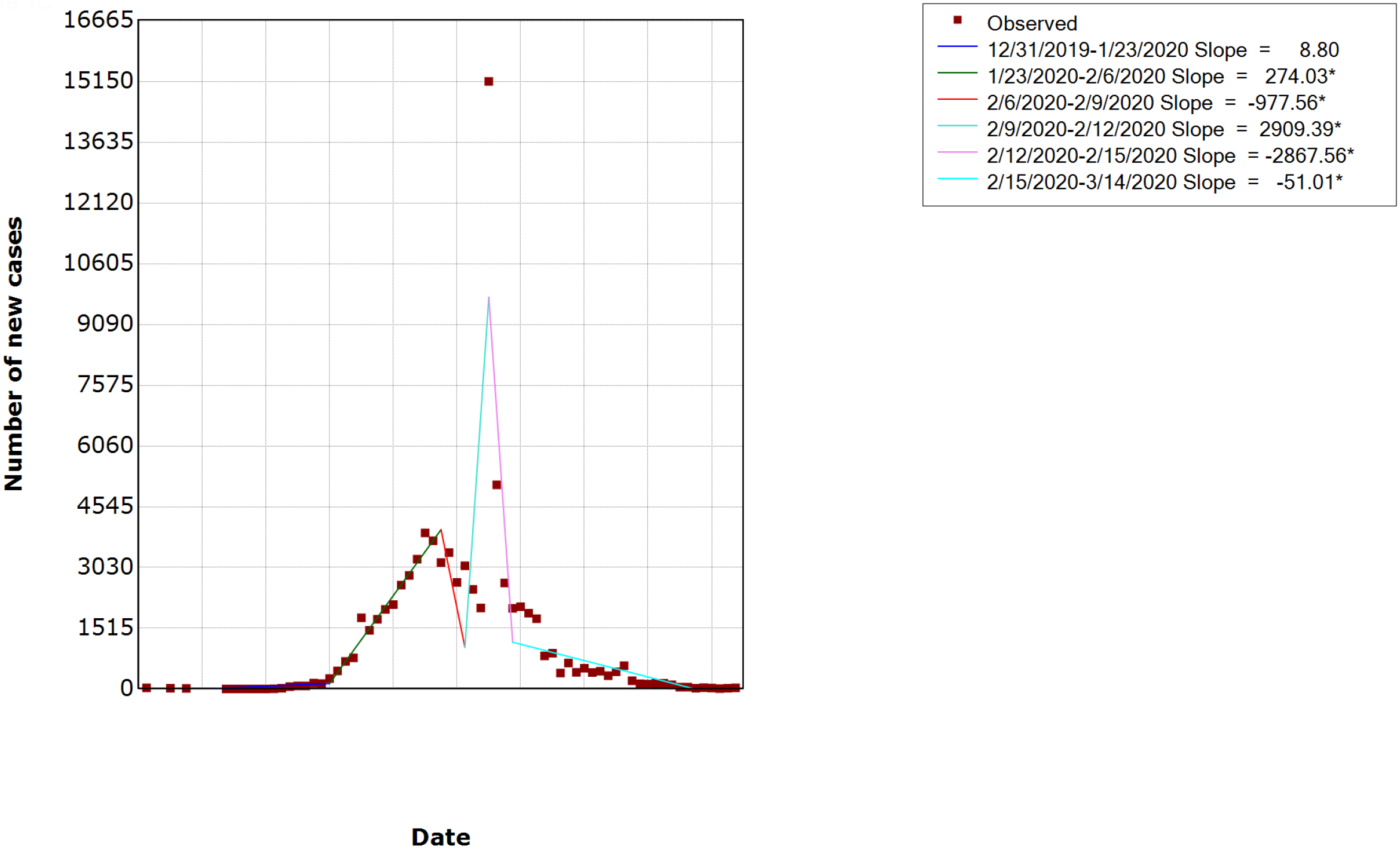
Jointpoint Regressions for COVID-19 cases reported between 31 Dec 2019 and 14 Mar 2020. Figure 1A: Number of new cases reported outside China; Figure 1B: Number of cumulative cases reported outside China; Figure 1C: Number of new cases reported in China * Indicates that the Slope is significantly different from zero at alpha=0.05 level

A similar growing trend was observed for cumulative cases although more exaggerated in the world outside of China. The trend was gentle at the first stage (13^th^ Jan. – 18^th^ Feb., slope = 18□5) with the number of cumulative cases slowly raising for about one month till it reached 804 on 18^th^ Feb. At the second stage (18^th^ – 27^th^ Feb., slope = 278□3), the cumulative cases increased to 3,664 in ten days and rapidly grew to 14,768 in about one week at the third stage (27^th^ Feb. – 5^th^ Mar., slope = 1549□0). The rapid increase continued to accelerate at the fourth stage (5^th^ – 11^th^ Mar., slope = 3756□2) and the fifth stage (11^th^ – 14^th^ Mar., slope = 8122□6). The cumulative cases had more than doubled in only one week and reached 37,371 as at 11^th^ Mar. at the fourth stage and climbed to 61,518 on 14^th^ Mar. at the fifth stage. This sharp increase in number of new cases and cumulative cases indicated that the world is experiencing a severe epidemic outbreak of COVID-19 with little evidence of mitigation.

Along with the increasing trend of new and cumulative cases, the number of countries and territories outside of China with reported COVID-19 cases also increased from one on 13^th^ Jan to 134 on 8^th^ March (Figure 2). Among these 134 affected countries and territories, 59 reported no more than 10 cases, 49 reported over 10 cases, 18 reported over 100 cases, six reported over 1000 cases, and two reported over 10,000 by 8^th^ Mar. Overall, the number of affected countries and territories experienced two increases. The first increase started from 23^rd^ Jan till 1^st^ Feb., where most countries reported 10 or fewer cases while several countries reported over 10 cases from 28^th^ Jan. The increase in number of affected countries then slowed and only five new countries reported COVID-19 cases from 1^st^ – 23^rd^ Feb., leading to a total of 28 affected countries by 23^rd^ Feb. However, the confirmed cases continued to increase in some affected countries as number of countries with over 10 reported cases grew from 5 to 10 on 1^st^ Feb, and South Korea, Japan, and Italy reported over 100 cases on 24^th^ Feb. The second apparent increase in the number of affected countries started from 25^th^ Feb. till 8^th^ Mar, during which 101 more countries reported confirmed COVID-19 cases. South Korea was the first country outside of China that reported over 1000 cases on 26^th^ Feb, but the number of its total confirmed cases was then surpassed by Italy, which became the first country with confirmed cases exceeded 10,000 on 11^th^ Mar. Six countries (Korea, France, Spain, Germany, United States, and Switzerland) had reported over 1,000 cases, among which four were European countries. Especially, the areas surrounding Italy has become a new large epicenter, in where the number of newly confirmed cases increases very rapid. More and more countries had reported over 10 or 100 cases during the second increasing period, although countries with less than 10 cases remained the majority. The fast increase of infected countries and confirmed cases demonstrate a widespread of COVID-19 globally and the occurrence of epidemics in regions outside of China.

**Figure 2:**
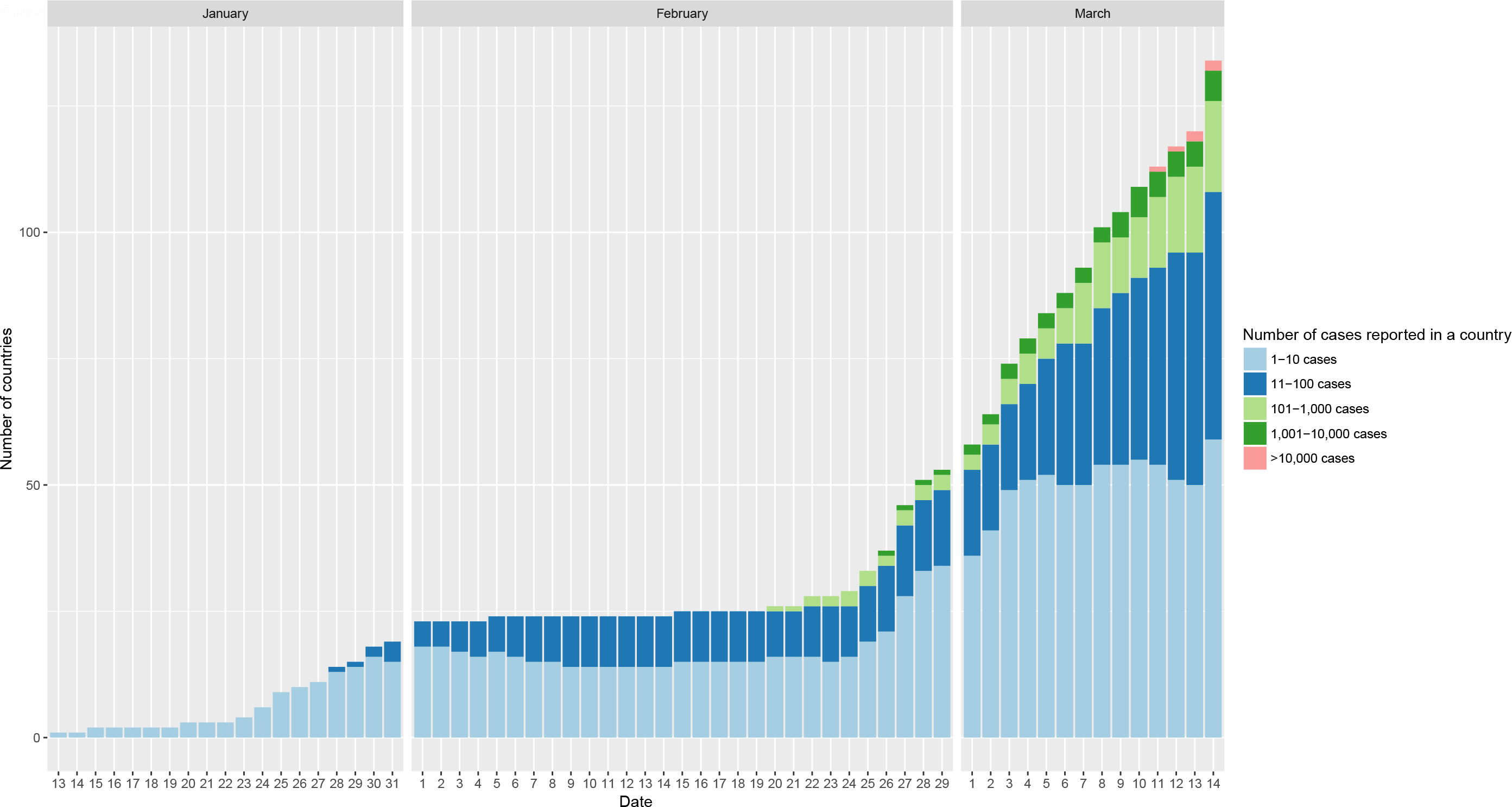
Number of infected countries in the regions outside China, 13 Jan – 14 Mar, 2020.

### Global transmission of COVID-19 outside of China

The first COVID-19 cases in different countries was captured on three maps: 28^th^ Feb. (the date when WHO first classified the transmission type for affected countries), 7^th^ Mar., and 14^th^ Mar. (Figure 3). The maps showed that Italy and Iran were the new epidemic centers for exporting COVID-19 as majority of the newly affected countries were close to and reported their first cases originated from the two countries (Figure 3 and Table S1). However, many neighboring countries and territories of Italy were affected and have developed local transmission compared to those of Iran, implying a faster spread of the disease in Europe than the Eastern-Mediterranean Region. For the affected countries in Africa and Latin America, many of them started to report COVID-19 cases in the week of 8^th^ −14^th^ Mar., when the disease spread quickly to the world and became a global pandemic. During this period, the first cases reported by the newly affected countries in Latin America and Africa were mainly imported from European countries, especially from Italy and France. The world was in a severe situation of the COVID-19 outbreak.

**Figure 3:**
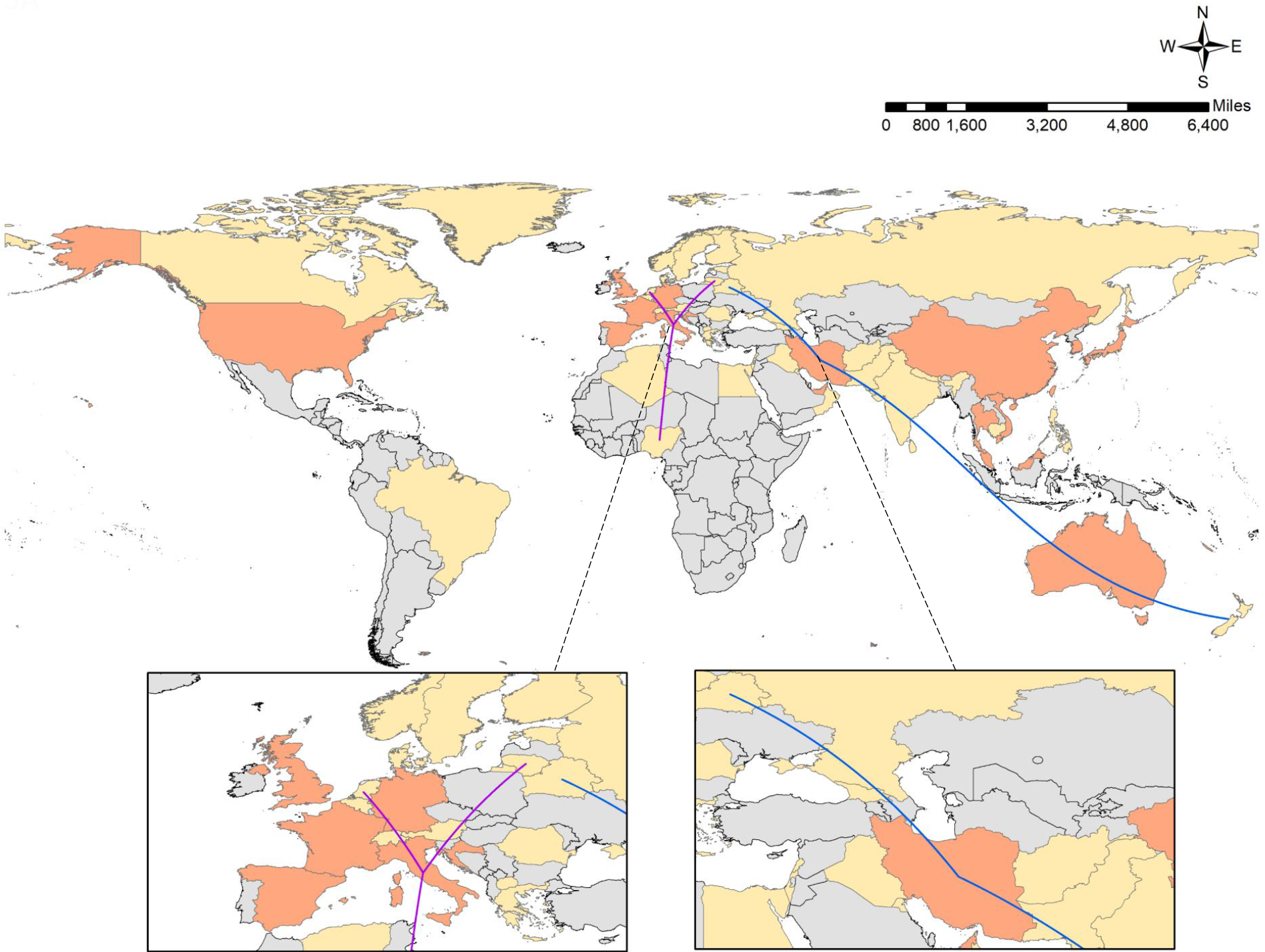
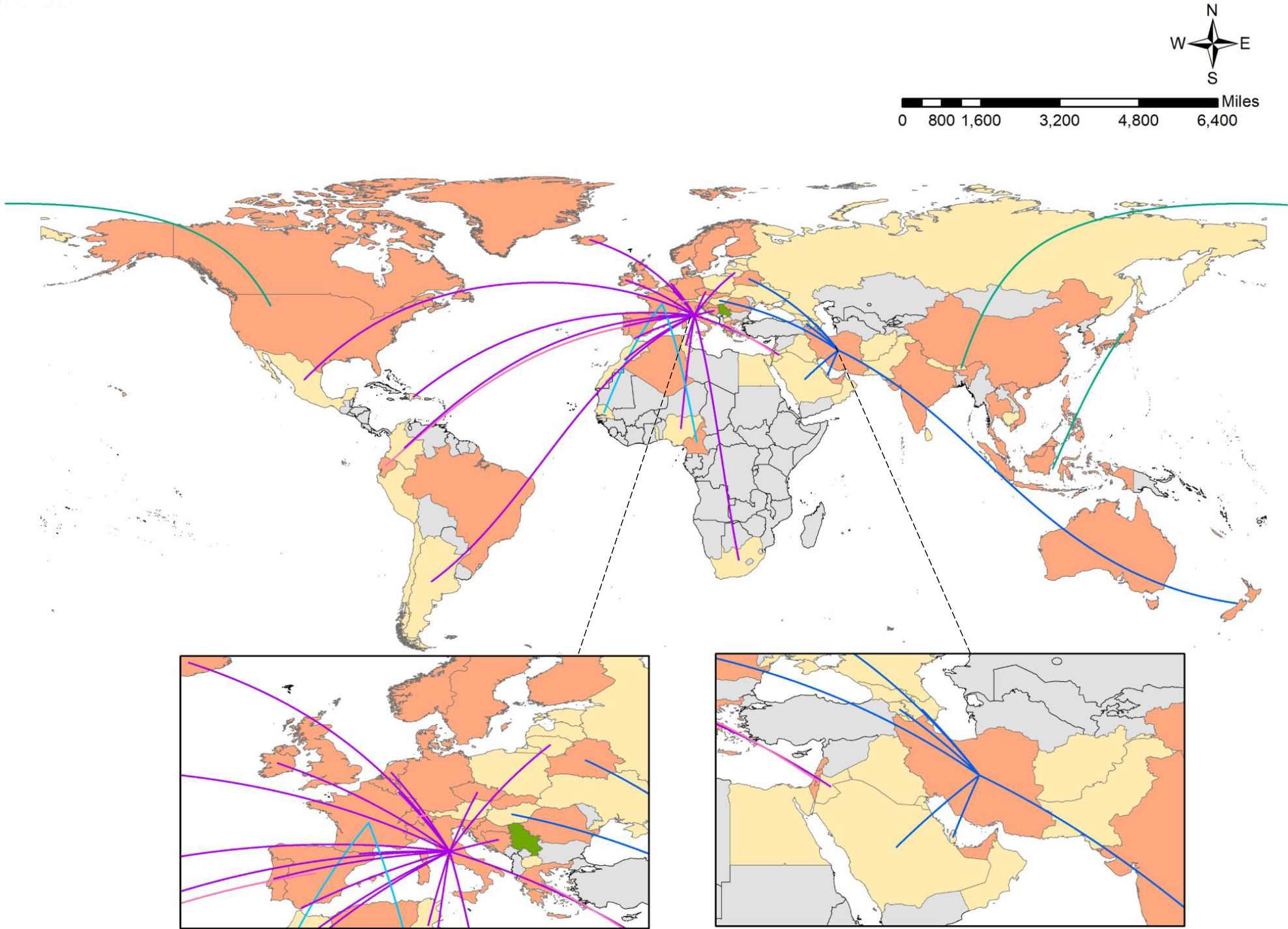
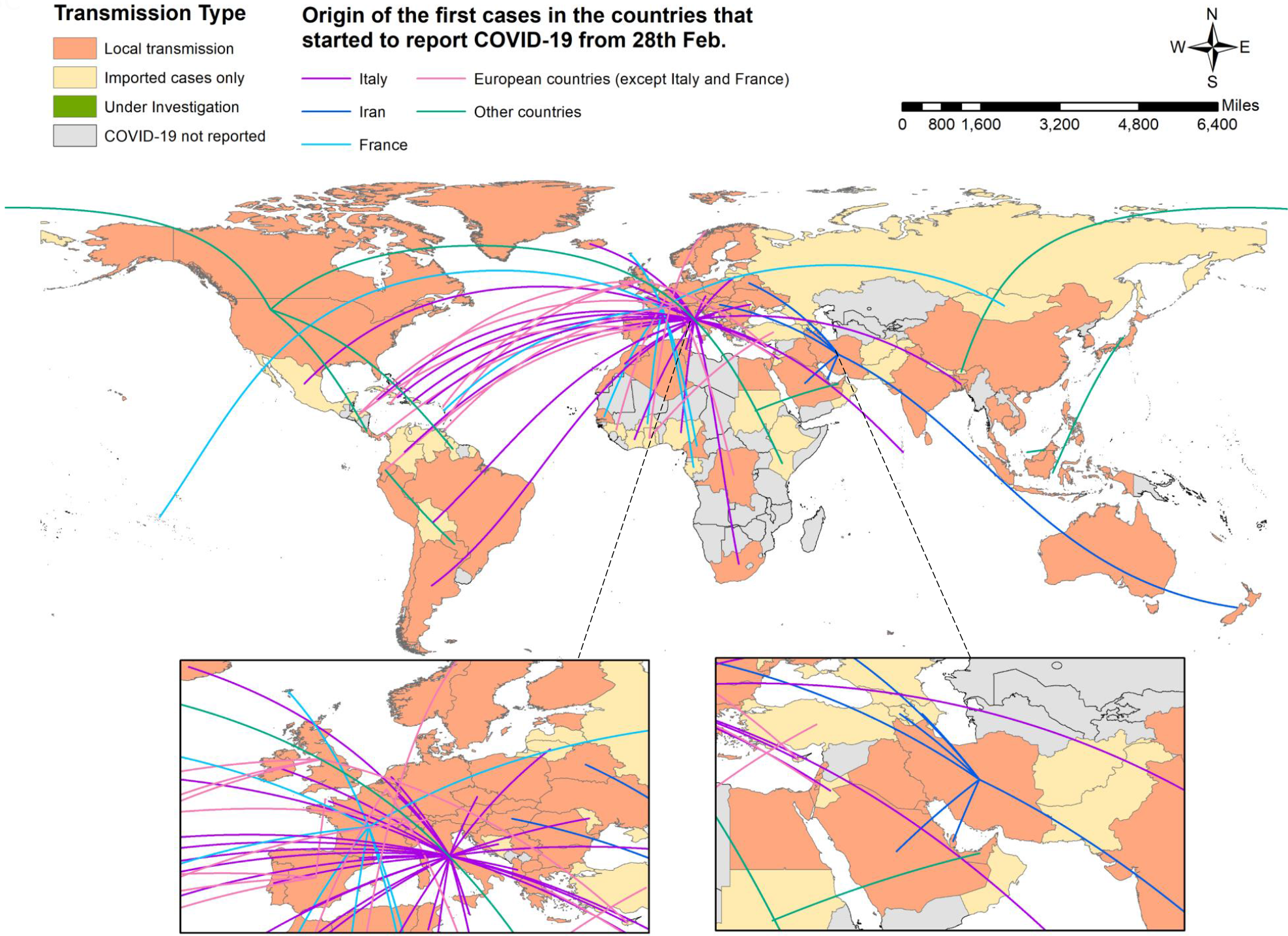
Global transmission maps of first reported cases by country in the regions outside China, 2020*. Figure 3A: Global transmission map of first reported cases by 28^th^ Feb.; Figure 3B: Global transmission map of first reported cases by 7^th^ Mar.; Figure 3C: Global transmission map of first reported cases by 14^th^ Mar. *Transmission classification was defined and classified by WHO.^1^ “Local transmission” refers to the phenomenon of local infection. “Imported cases only” refers to the phenomenon of patients infected outside the reported location. “Under investigation” refers to the situation that the transmission type of the location was unknown.

### Geographic distribution of COVID-19 cases

Dynamic maps of the disease distribution were produced to show the changes in new cases and cumulative cases in the world during the COVID-19 outbreak (Supplement 2 and 3). In general, the outbreak can be divided into two stages: a single-center period and a multi-centers period. During the single-center period (13^th^ Jan. - 20^th^ Feb.), the epidemics mainly occurred in China, which had 75,993 confirmed cases while other countries had around or less than 100 cases by 20^th^ Feb. Most affected countries were from Europe and the Western Pacific Region while no case was reported in South America within this period.

During the multi-centers period (after 20^th^ Feb.), the number of new cases gradually decreased in China while rapidly increased in South Korea from 20^th^ Feb., in Iran from 22^nd^ Feb., and in Italy from 23^rd^ Feb. The disease also spread to many other countries and territories in Europe and the Eastern Mediterranean Region as well as the countries and territories in Africa and South America, which were the two continents with fewer infections in the single-center period. South Korea, Italy, and Iran became the new centers of the outbreak as of 8^th^ Mar, with Italy and Iran reporting over 10,000 cases and South Korea reporting over 8,000 cases. France, Spain, Germany, Switzerland, and United States became minor centers of the epidemic with over 1,000 cases reported.

## Discussion

Sustained human-to-human transmission of the COVID-19 worldwide appears inevitable and understanding the real-time patterns of the epidemic would be useful for guiding further time-sensitive responses. This study estimated the trend, the transmission patterns, and the geographic distribution of the COVID-19 global epidemic from 31^st^ Dec. 2019 to 14^th^ Mar. 2020. We found a multi-centers pandemic has been established. This study adds to the existing report of the WHO and literature by systematically evaluating the global changing patterns of the epidemic, comparing this change with China, and evaluating the geo-spatial pattern of the pandemic.

We found the sharp increasing trends in new cases and cumulative cases and the expanding number of epidemic centers, indicating that the world is experiencing a severe COVID-19 pandemic with little evidence of mitigation. The tremendous growth of new cases indicates sustained human-to-human transmission has been established globally although the epidemic is under control in China. In response to the epidemic, countries should not only follow the Public Health Emergency of International Concern (PHEIC) issued by WHO, and take effective actions to halt the social distance;[11] but also fully expand the screening, diagnosis, and treatment of the existing cases, strengthening quarantine and isolation strategies; as well as expand the supply of medical supplies, including personal protective equipment and lab testing materials. In addition, countries in Africa and Americas should try their best to avoid becoming the next epicenter.

Different epidemiological patterns of the COVID-19 epidemic were observed between China and the world outside of China. Our findings showed that countries outside of China experienced a much longer slow-growing stage but long increasing period, and it is hard to predict when the global epidemic will reach a turning point. These results have several important implications: First, the world lost the most valuable time for COVID-19 control after the outbreak of COVID-19 in China. After China taking the comprehensive prevention and control strategies, and after the WHO issued the PHEIC, many countries did not take enough precautionary actions to prevent the establishment of local transmission and became the new epidemic centers which further facilitated the pandemic. Second, the global situation is more complicated than that of China, and it is hard to take as the same level of action as China did. The global COVID-19 epidemic is under the impact of available of medical resources, religion, political, economic and other comprehensive reasons, and as these vary from country by country, a “one-to-all” strategy may not suit all. Thus, tailored strategies are needed for each infected country, based on the maximum resources they have, while social distance keeping is one of the most powerful and cost-effective approach. Third, even the Africa region and other low- and middle-income countries (LMICs) are currently only slightly under the impact of the epidemic, 15 countries already have confirmed cases. Thus, restrict reactions are should be further strengthen in this area. Limited resources for COVID-19 screening and testing may have the COVID-19 epidemic underestimated in LMIC settings, and not taking enough attention for the further control of the epidemic may brew another tsunami which will further worsen the pandemic.[12] LMIC countries should fully implement robust containment and control activities as suggested by WHO and evidenced by the Chinese experience.[13]

Our study indicated that local transmission has been established in more than 50% of the countries that reported cases of COVID-19. The establishment of local transmission and the increase in the number of epidemic centers indicates a widespread and that a sustained transmission of COVID-19 has been established globally, and therefore, the war against COVID-19 may have changed from an emerging response to a long-term war. Thus, facilitating the development of rapid, cheap and high accuracy testing methods, accelerating the development of treatment and vaccines, increasing the industries’ ability in medical supplies manufacturing, and improving the access to medical care are essentially urgent in preparing for this long-term battle.

This study has some limitations. First, the data collected were from publicly available datasets. They lacked detailed epidemiological information as well as the detailed geographic information on patients for further assessing the potential driving forces as well as more detailed geographic patterns of the pandemic. Secondly, our analysis is lagged by a time gap between when a suspected case identification and case confirmation, but it does provide time-sensitive and evidence-based information to aid in further response to the epidemic. Finally, due to constraints of detection capability and technique, the underreporting of cases especially in developing countries may result in underestimation of the study.

## Data Availability

All the data are publicly available at World Health Organization, China's National Health Commissions, and GADM.org

https://www.who.int/emergencies/diseases/novel-coronavirus-2019/situation-reports

https://gadm.org/download_country_v3.html

http://www.nhc.gov.cn/xcs/yqtb/list_gzbd.shtml

## Reference

1. WHO. Novel Coronavirus (2019-nCoV) situation reports: World Health Organization; 2020 [Cited 14 March 2020]. Available from: https://www.who.int/emergencies/diseases/novel-coronavirus-2019/situation-reports

2. WHO. Joint WHO and ECDC mission in Italy to support COVID-19 control and prevention efforts. 2020 [Cited 14 March 2020]. Available from: http://www.euro.who.int/en/health-topics/health-emergencies/coronavirus-covid-19/news/news/2020/2/joint-who-and-ecdc-mission-in-italy-to-support-covid-19-control-and-prevention-efforts.

3. WHO. Coronavirus disease 2019 (COVID-19) Weekly situation report, Issue 1. 2020 [Cited 14 March 2020]. Available from: http://www.emro.who.int/images/stories/coronavirus/covid-sitrep-issue1.pdf?ua=1.

4. WHO. WHO Updated Country Preparedness and Response Status for COVID-19 as of 6 March 2020: World Health Organization; 2020. 2020 [Cited 14 March 2020]. Available from: https://www.who.int/internal-publications-detail/updated-country-preparedness-and-response-status-for-covid-19-as-of-6-march-2020.

5. The Times of Israel. Iraq closes border with Iran over Coronavirus fears. 2020 [Cited 14 March 2020]. Available from: https://www.timesofisrael.com/iraq-closes-border-with-iran-over-coronavirus-fears/.

6. BBC News. Countries close borders as Coronavirus spreads. 2020 [Cited 14 March 2020]. Available from: https://www.msn.com/en-xl/news/other/countries-close-borders-as-coronavirus-spreads/ar-BBZxY2v.

7. Dormido H, Leung A. Charting the global economic impact of the Coronavirus. 2020 [Cited 14. March 2020]. Available from: https://tbsnews.net/international/global-economy/charting-global-economic-impact-coronavirus-42439.

8. Gill A. The Impact of the Coronavirus on Global Economy. 2020 [Cited 14 March 2020]. Available from: https://www.globalbankingandfinance.com/the-impact-of-the-coronavirus-on-global-economy/.

9. WHO. Shortage of personal protective equipment endangering health workers worldwide. World Health Organization, 2020 [Cited 14 March 2020]. Available from: https://www.who.int/news-room/detail/03-03-2020-shortage-of-personal-protective-equipment-endangering-health-workers-worldwide.)

10. GADM. GADM maps and data. Oxford: GADM; 2020 [Cited 14 March 2020]. Available from: https://gadm.org/download_country_v3.html

11. Anderson RM, Heesterbeek H, Klinkenberg D, Hollingsworth TD. How will country-based mitigation measures influence the course of the COVID-19 epidemic? The Lancet 2020. doi:10.1016/S0140-6736(20)30567-5

12. Ebrahim SH, Memish ZA. COVID-19: preparing for superspreader potential among Umrah pilgrims to Saudi Arabia. The Lancet 2020.

13. Zhang S, Diao MY, Duan L, Lin Z, Chen D. The novel coronavirus (SARS-CoV-2) infections in China: prevention, control and challenges. Intensive Care Medicine 2020:1–3.

